# COVID-19 Spread in India: Dynamics, Modeling, and Future Projections

**DOI:** 10.1101/2020.06.12.20129197

**Authors:** Rajesh Ranjan

## Abstract

COVID-19 is an extremely infectious disease with a relatively large virus incubation period in the affected people who may be asymptomatic. Therefore, to reduce the transmission of this pathogen, several countries have taken many intervention measures. In this paper, we show that the impact of these measures in India is different from several other countries. It is shown that an early lockdown in late March 2020 changed the initial exponential growth curve of COVID-19 to a linear one, but a surge in the number of cases from late April 2020 brought India back to a quadratic trajectory. A regional analysis shows the disparate impact of the intervention in different states. It is further shown that the number of reported infections correlates with the number of tests, and therefore regions with limited diagnostics resources may not have a realistic estimate of the virus spread. This insufficiency of diagnostic test data is also reflected in an increasing positivity rate for India nearly 2.5 months after the lockdown, inconsistent with the trends observed for other geographical regions. Nonetheless, future projections are made using different epidemiological models based on the available data, and a comparative study is presented. In the absence of a reliable estimate of the true number of infections, these projections will have a limited accuracy: with that limitation, the most optimistic prediction suggests a continuing virus transmission through September 2020.

## 1 Introduction

Coronavirus disease 2019 (COVID-19), a respiratory disease caused by the SARS-CoV-2 virus, has been declared as a pandemic by the world health organization (WHO) in March 2020. The severity of the virus spread can be understood by looking at the mortality figures, despite a lower case fatality rate (CFR) than that of SARS [1]. As of June 10, 2020, this disease has claimed about 0.42 million lives worldwide, correspondingly with about 7.4 million confirmed positive cases.

The first COVID-19 case was detected in late December 2019 in the Wuhan province of China. After widespread transmission in China, the disease spread to the European Union (EU) nations, followed by a major outbreak in the United States (US). In recent days, a steep increase in cases is noticed in India, Russia, and South American nations. In the absence of any available vaccine for COVID-19, countries worldwide have implemented numerous social distancing measures to limit its transmission and control the outbreak as well as to delay the peak. Despite that, the rapid rise in the number of cases has put a significant strain in the healthcare systems.

The present study is focused on the characterization of COVID-19 spread in India: the impact of lockdown on the growth curves, and mathematical models to study the spatio-temporal dynamics based on data till June 10, 2020. For future references, 2020 is the default year for all the dates referred, unless mentioned otherwise. India first reported a COVID-19 case in a student who returned from Wuhan, China on January 30, 2020. Since then, there has been a rise in the number of infections with 0.28 million cases on June 10, among which only 50% are active cases and rest are either recovered or deceased. CFR for India is relatively low with total deceased cases is 0.28% of the total cases. High recovery rate and low CFR may be due to the factors like a large proportion of the young population, and possible immunity due to BCG vaccinations[2]. However, most of these studies are preliminary and correlation-based, and therefore more evidence is required for arriving at concrete conclusions[3].

The spread of COVID-19 in India is compared against the most affected countries as well as China and South Korea in Fig. 1(a,b). The time-series data has been taken from ‘Our World in Data’ (OWID) source that compiles data from the European Centre for Disease Prevention and Control (ECDC). In Fig. 1(a), we notice dissimilar curves for virus spread in different countries. The curves for China and South Korea are completely flattened, who used different approaches to isolate the cases with virus infections and thus successfully controlled the spread. Most of the EU nations, after a prolonged nearly linear growth, are now showing signs of flattening. For both US and Russia, the curves represent linear growth, while for India and Brazil the curves can be best described by an exponential or a power law.

**Figure 1:**
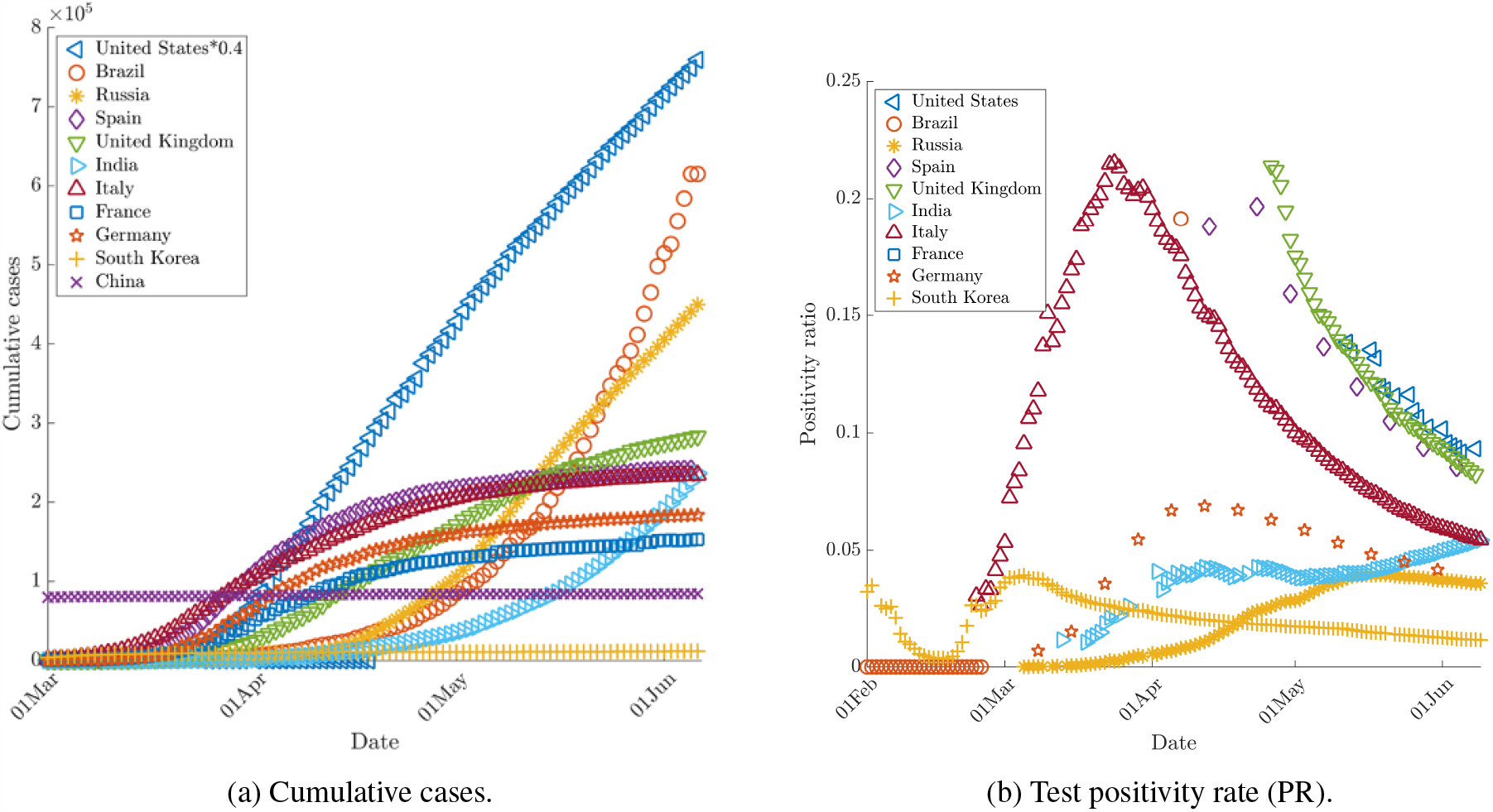
Comparison of Covid-19 spread in India with several other key countries.

An important indicator that provides insights on the dynamics of COVID-19 spread is the cumulative test positivity rate (PR), defined as the total number of positive cases per total number of tests. An increase in this number indicates that the severity of spread is increasing, and testing capacity needs to be ramped up for the size of the outbreak to keep this number low. India has relatively low PR (5.7%) compared some of the worst affected countries like Brazil and the US which has PR up to 38% and 13% respectively on June 8. Fig. 1(b) shows the temporal variation of these ratios for these countries, as available from OWID source. Most of the regions shown in this figure, exhibit a bell-shaped curve with PR increasing initially as the spread intensifies faster than the rate of increase in the testing capacity, and then a decay due to interventions as well as increased rate of tests per capita. All the countries in this figure, except India, show a decline in PR in recent days. This inconsistent increasing trajectory for India 2.5 months after the lockdown could be ascribed to insufficient testing capacity[4]. As of June 8, 2020, India has about 3,400 tests per million inhabitants, compared to 66,000 and 90,000 tests per million for the US and Russia. Reference[4] have recommended that in order to have a reliable number of total infections and keep the PR to the recommended level of 5% or below, India’s testing capacity must be escalated to about 1 million per day compared to current capacity (on June 8) of about 0.15 million per day.

The dynamics of COVID-19 growth in India is slightly unanticipated as India implemented international travel bans and took strict social distancing measures quite early on March 23, when the number of cases was very low (536). In order to assess the impact of lockdown in India compared to other nations, we show their growth curves before and after lockdown on the same scale as shown in Fig. 2(a). In this figure, day 0 (*t*_0_) shows the cases when social distancing measures were implemented. The orange shaded region, between *t*_∗1_ and *t*_0_ shows the continuing initial exponential growth typical of an epidemic, which continues through 14 days after the lockdown date to *t*_1_ = *t*_0_ + 14. The grey shaded region indicates the days from *t*_1_ when the effects of social distancing are expected to appear.

**Figure 2:**
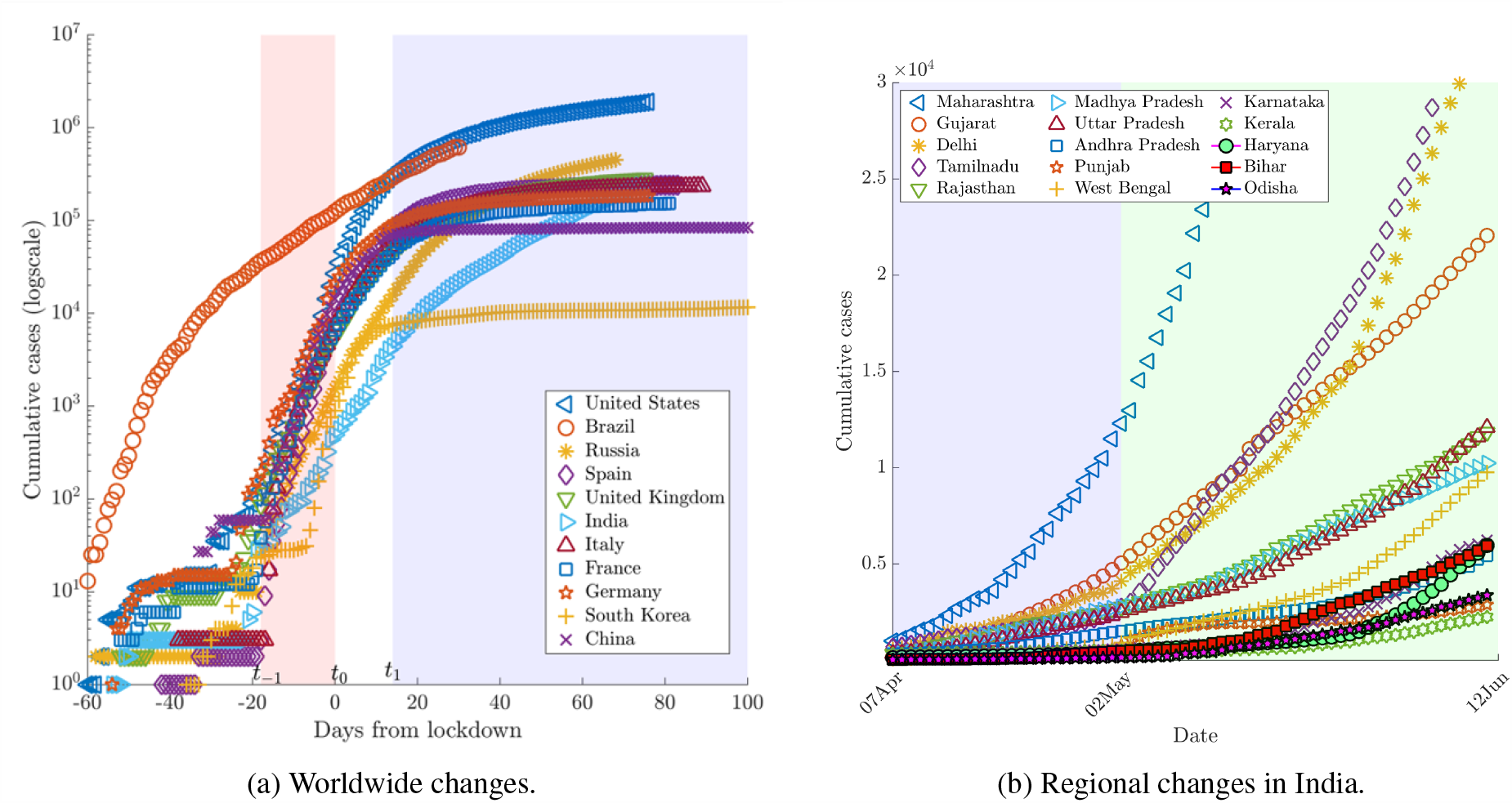
Change in growth curve due to lockdown measures.

Unlike China and South Korea that show rapid flattening of the curves soon after *t*_1_, other countries exhibit only a slow down in the growth rate. The signs of flattening for European countries are observed about a month from *t*_1_. This phase has been described by power laws [5] or long linear regime [6]. The curves for the US and Russia show slow flattening. In Brazil, the lockdown measures were taken very late in early May and therefore the signs of flattening are still not visible. The case of India is surprising as the curve continues to rise even after 80 days from the date when lockdown measures were implemented.

In order to further analyze this unanticipated trend for India, we focus on the regional dynamics. Figure 2(b) shows the curves post-lockdown for a few key states. The trends observed for different states are very dissimilar, which may indicate different compliance levels of lockdown measures in these regions. According to an estimate by Chang *et al*.[7] for the COVID-19 outbreak in Australia, a reduction in incidence and prevalence can be visible only if the social distancing compliance levels exceed 80%. While Kerala, which first reported COVID-19 infection in India, and several other states show a reasonable control of the pandemic, four states - Maharashtra, Delhi, Tamilnadu, and Gujarat continue to exhibit rapid growth, with the rate increasing in that order. This also highlights that a major contribution in the COVID-19 cases are due to urban areas i.e. Mumbai, New Delhi, Chennai and Ahmedabad. The figure also shows that, more recently, states like Bihar, Uttar Pradesh, Odisha, Rajasthan and West Bengal show a surge in the number of cases indicating that the virus may now be penetrating to the rural areas. This could be attributed to the movement of the migrant workers and laborers from the metro cities with clusters to the rural areas. The positivity rate among states also varies significantly. For example, on June 9, Karnataka has low PR for (1.48%), while Maharashtra (15.2%) and Delhi (12.3%) have high PR.

In this work, we study the changing COVID-19 dynamics in India presented above using various mathematical models. The growth of an epidemic is described by phenomenological or mechanistic models. Reference[8] has delineated popular phenomenological models used to estimate the initial growth of the epidemic, while the basic mechanistic compartmental models that characterizes the overall behavior from the beginning to end are detailed in [9]. An issue with an ongoing epidemic is that the epidemiological parameters such as infection and recovery rates, needed for the compartmental models are not precisely known. Therefore, these parameters are typically estimated using the available data by fitting the curves with different initial conditions to get the best fit. However, the temporal variation of these parameters due to social distancing measures or a change in testing criterion is not accounted in these models. Despite these limitations, these models provide valuable insights about the trajectory of the pandemic curve in a given region, if used with an understanding of the underlying assumptions. The dynamics of COVID-19 growth before and after the lockdown are discussed, and future projections are given.

## 2 Mathematical Models for COVID-19 Spread

In this work, we use four models, two phenomenological: the exponential and logistic, and two mechanistic: Susceptible-Infectious-Recovered (SIR) and generalized Susceptible-Exposed-Infectious-Recovered (SEIR) model, to describe the existing as well as project the future dynamics of COVID-19 in India. A description of these models is given in appendix A. Further, power-law models are used to describe post-lockdown dynamics. The spread dynamics are divided into two periods based on the chronology of COVID-19 spread, and pertinent models are used.

### 2.1 Early Growth and Post-lockdown Curves

As reviewed by Reference [8], the initial growth of an epidemic can be described by phenomenological models with any complication of parameter estimation in the mechanistic models. The exponential model describes the cumulative incidence curve of an epidemic reasonably well. In the present case, we use an exponential fit to describe the initial growth dynamics of COVID-19 in India.

Figure 3(a) shows the exponential fit obtained using the data between March 15 and March 29. Table 1 shows the estimated coefficients for this model. The lower and upper bounds at 95% confidence level are given, along with other statistical parameters such as standard error, t-stat and p-value for each coefficient. The statistical parameters of this entire regression model is given in Table 2. The coefficient of determination for this fit is *R*^2^ = 0.991. Further, a very small p-value (∗ 0.000) indicates that all the regression parameters are statistically significant.

**Table 1:** Exponential regression model for initial growth: *y* = *a*_1_ exp(*a*_2_*x*).

|  | Estimate | 95% CI |  | SE | tStat | pValue |
| --- | --- | --- | --- | --- | --- | --- |
| $a_1$ | 106.8 | 91.42 | 122.2 | 7.1312 | 14.975 | 1.4096e-09 |
| $a_2$ | 0.1607 | 0.1494 | 0.1719 | 0.0052211 | 30.771 | 1.5685e-13 |

**Table 2:** Statistics for Exponential, Logistic and Quadratic models

| Model | Exponential | Logistic | Quadratic |
| --- | --- | --- | --- |
| Number of observations | 15 | 15 | 45 |
| Degrees of freedom | 13 | 12 | 42 |
| Root Mean Squared Error | 32.9 | 19.9 | 1740 |
| R-Squared | 0.991 | 0.997 | 0.999 |
| Adjusted R-Squared | 0.991 | 0.997 | 0.999 |
| F-statistics vs. zero model | 2360 | 4320 | 98100 |
| p-value | 2.22e-17 | 1.84e-18 | 9.19e-81 |

**Figure 3:**
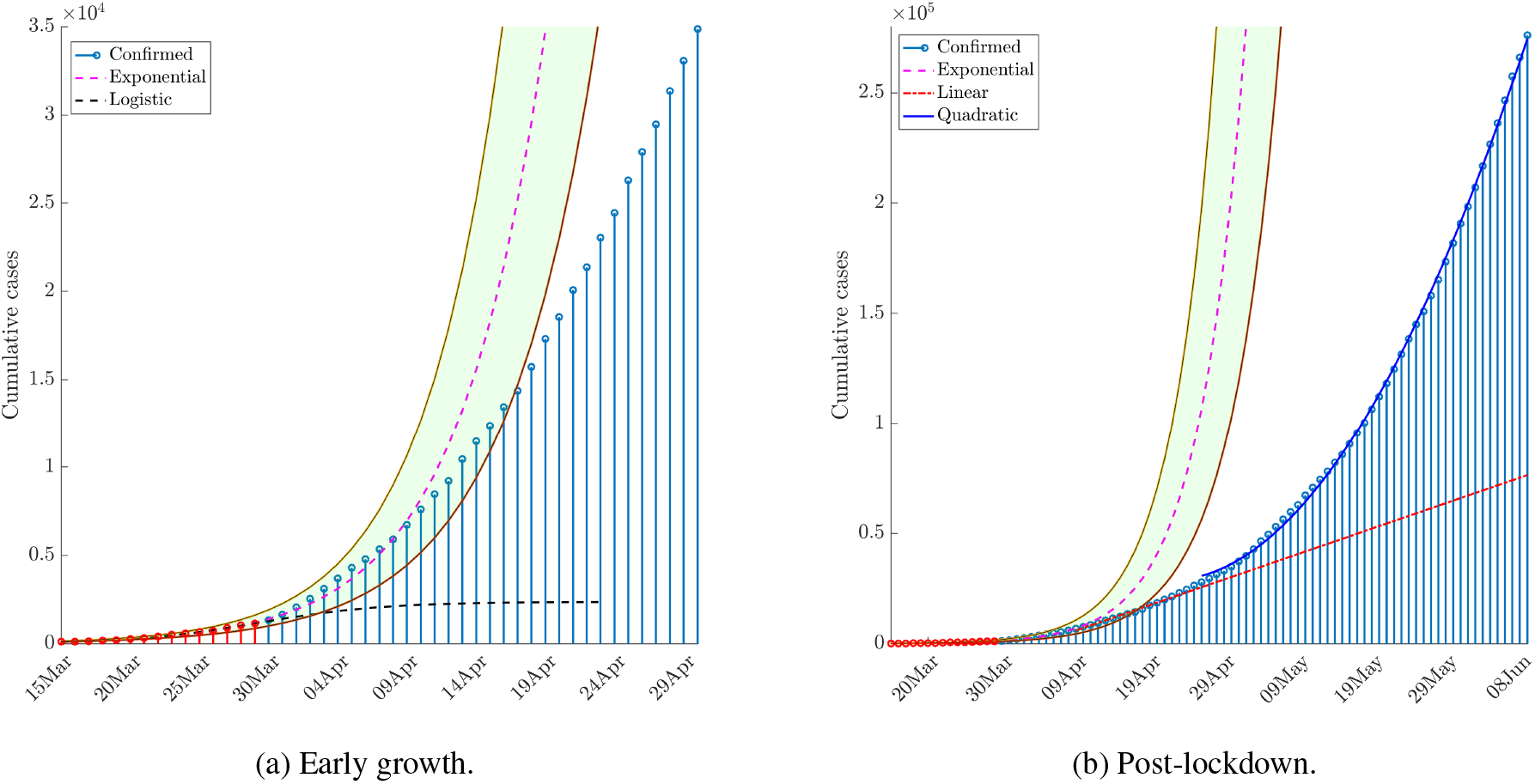
Early growth (from March 15 until April 9) follows the exponential model (a). Red stems show the data used for this exponential fit. Brief linear growth period between 9 Apr and 26 Apr is given by the curve *y* = 1154.7*x* + 4771.7(*R*^2^ = 0.9898, RMSE = 505.48. Since late April, the cases exhibit a quadratic growth curve (b). Coefficients and statistical parameters for exponential and quadratic curves are listed in Tables 1,2 and 3

This exponential model predicts the cases reasonably well until April 9 as shown in the figure. It is to be noted that India implemented the lockdown measures on March 23, so the exponential growth continues for about 15 days after this based on the virus residency period before we note a systematic departure from the exponential curve. For comparison, a logistic fit (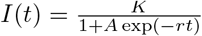, with *K* = 2368, *r* = 0.22087, *A* = 23.695) using the same dataset, is also shown in the same figure. This model predicts flattening soon after April 3, highlighting the limitation of using this model with an insufficient dataset.

Now, the post-lockdown dynamics of COVID-19 are described. Figure 3(b) shows the growth curves in this phase. Immediately after the lockdown, the cumulative growth trend shifts to a linear curve as shown by the red solid line. This continues for 15 days between April 10 and April 25, with about an average of 2000 cases per day. However, after this a systematic departure from the linear curve begins, and the growth now exhibits a power-law as suggested by Reference [10]. A fit using the data between April 25 and June 10 shows a quadratic growth curve, with their coefficients given in Table 3. Statistical parameters for this fit shown in Table 2 shows a good significance.

**Table 3:**
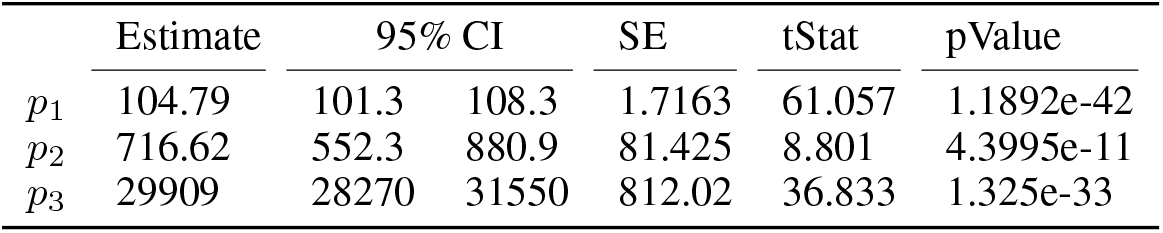
Quadratic regression model post-lockdown: y = p1x^2^ + p2x + p3.

The shift from linear to quadratic growth during the lockdown is not anticipated. In order to elucidate this, we compare the diagnostic data with the number of infections as shown in Fig. 4(a,b). On the logscale shown in Fig. 4(a), we note that number of daily infections is proportional to the number of tests conducted daily. So, the short linear growth period with relatively post-lockdown could be attributed to an insufficient number of tests during this period. For comparison, the number of daily tests conducted on April 9, April 29 and June 7 is 16,991, 54,031, and 1,42,069 respectively. Apart from an order of magnitude in the testing capacity compared to that in early April, the recent rapid growth could also be due to the inclusion of asymptomatic patients in the testing criterion.

**Figure 4:**
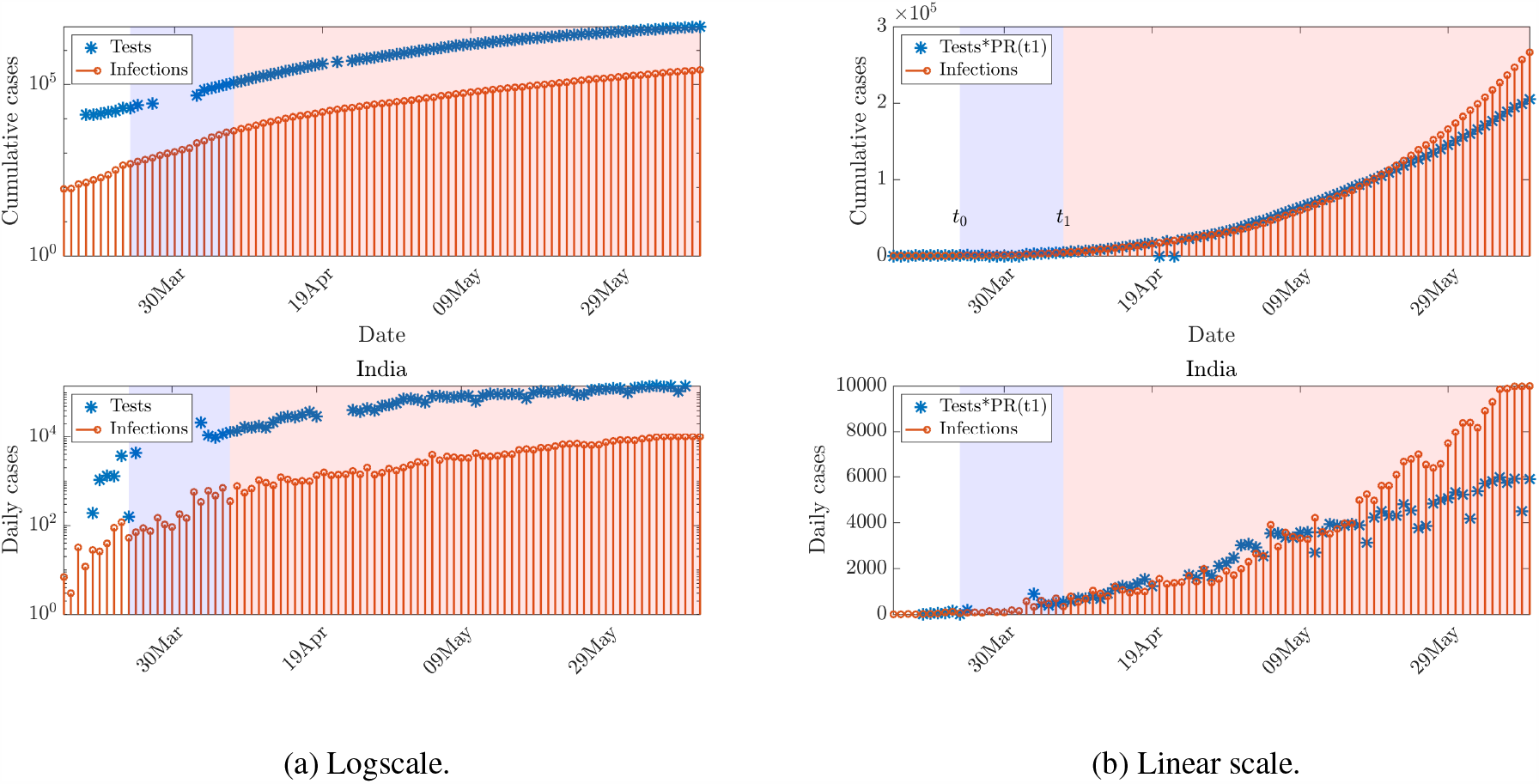
Comparison of Covid-19 growth in India with diagnostic test data. The shaded region show the period from implementation of lockdown (*t*_0_) to *t*_1_ = *t*_0_ + 14.

A plot on the linear scale shown in Fig. 4(b) provides further insights. The test data on the *y*-axis is normalized with positivity rate (PR) on day *t*_1_ (14 days from the lockdown date; PR(*t*_1_)=0.0417) as defined in Fig. 2(a). It is expected that if the lockdown is effective and the testing criterion includes sufficient susceptible population, PR(*t*) should decrease from PR(*t*_1_) value regardless of change in testing capacity as seen from the bell-curves in Fig. 1(b) for several other countries. Therefore, the time curves of infections should go below that normalized test (tests × PR(*t*_1_)) curve with time. However, for India, a reverse trend is obtained as shown in Fig. 4(b) for both daily as well as cumulative cases. In both plots, the infection curve is consistently above the normalized test curve from the middle of May, This unexpected increase in total infections per tests, attributable to insufficient testing capacity as argued by Reference [4], is an important factor for the quadratic growth in the number of cases shown in Fig. 3(b).

### 2.2 Peak and Subsequent Decay

In this section, we use both phenomenological (logistic) and mechanistic (SIR and SEIQRDP) models to predict the peak ans subsequent decay of COVID-19 spread in India. The underlying epidemiological parameters for SIR and SEIQRDP models are obtained by first making an initial guess, and then minimzing the error between the results from the model and the actual data in a least-squares sense. Open-source MATLAB codes developed by Batista[11] and Cheynet[12] are respectively used for SIR and SEIQRDP models. These codes use different initial guess for estimating the epidemiological parameters, but the predictions by both these models are similar if sufficiently long data in the decay phase are available[13]. On the other hand, there could be a high uncertainty in the prediction of peak[14, 15].

Figure 5 shows these predictions from logistic, SIR, and SEIQRDP models for India based on data until June 8. As anticipated, the prediction of peak from all these models is very different. The logistic model predicts the peak quite early soon after the timeline data, while the SEIQRDP model predicts the peak to be in the middle of July with about 17,000 cases per day. Because of this, there is a high variation in the estimated cumulative number of cases by these models. Logistic, SIR and SEIQRDP models respectively project 0.6, 1.0 and 1.9 million cases by the middle of September. The growth estimate obtained by extrapolating the current quadratic trend given by table 3 is also shown. If the current trend continues, India will have over 1 million cases by the end of July.

**Figure 5:**
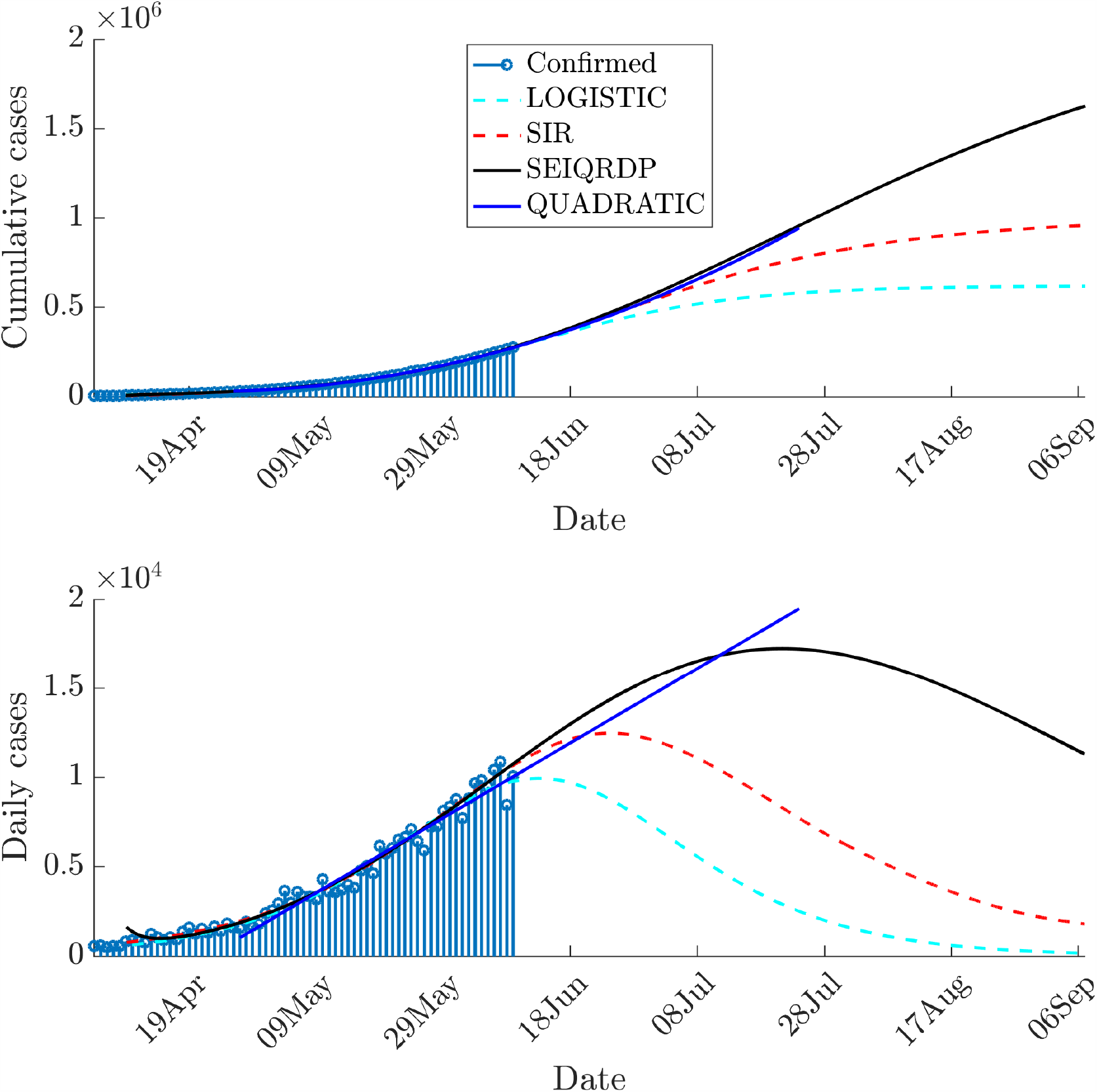
Projected trajectory for COVID-19 based on mechanistic and phenomenological models.

As seen from the recent trends from European nations as well as the US, the decay in the number of cases after the peak is very slow compared to its growth before the peak (see Fig. 1(a)). Therefore, it is unlikely that during the decline phase, India will follow the fast decay curve predicted by the models shown in Fig. 5 and a more reliable data-driven approach should be used in this phase [15]. Considering these factors, it is implausible that a complete flattening of COVID-19 curve will be achieved before October even in the most optimistic case. However, an excellent recovery rate for India combined with a low case fatality rate are some favorable signs that suggest that the social impact due to this spread may still be relatively less devastating. Further, these models do not account for a possible change in infectivity of SARS-CoV-2 due to changes in Meteorological parameters such as a variation in temperature or humidity[16] or air index[17].

Lastly, as discussed earlier in Fig. 2(b), the dynamics of COVID-19 spread in different states are very dissimilar. Therefore, it may be more dependable to use these models for statewise predictions. However, one of the primary assumptions of these models is that the considered population should not change due to immigration. Because of the large movement of migrant workers and laborers, which constitute a susceptible population, this assumption may become invalid when states are modeled separately.

## 3 Conclusions

In this work, the dynamics of COVID-19 spread in India from the beginning until June 8, 2020 are analyzed. Different mathematical models are used to describe the initial rapid growth (phase-1), slow growth immediately after lockdown (phase-2), and later and more recent fast growth (phase-3). It is shown that exponential, linear and quadratic curves define these phases with high statistical significance. The switch from linear to quadratic growth after lockdown is suspected to be related to the order-of-magnitude increase in the diagnostic testing capacity. However, an increase in the positivity rate nearly 1.5 months after the lockdown, inconsistent with what is observed for other countries, indicates that the testing capacity may still not be adequate. Future projections based on available data are made using both phenomenological and mechanistic models. These models have high variability in the prediction of the peak, and even the most optimistic case projects the flattening of the curve in September. The accuracies of these predictions are also limited in the absence of an accurate estimate of the true number of cases due to insufficient testing capacity. Therefore, these models should be regularly revisited, as more and more reliable estimates of the infections become available with increasing testing capacity. The variability in estimates from different models will also decrease once India enters the decline phase.

## Data Availability

The data included in the study are publicly available.

https://ourworldindata.org/

## A Description of Mathematical Models

### A.1 Exponential Model

In this model, the number of diagnosed infections *I*(*t*) increase exponentially with time,

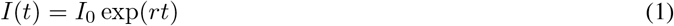

Where *I*_0_, *r* are the initial number of infections and growth rate respectively.

### A.2 Logistic Model

The the growth of the pandemic using a logistic model is given by[18]:

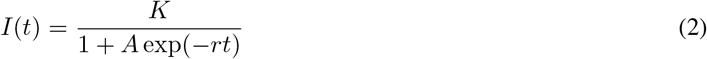

Where *A*= (*K*/*I*_0_) − 1 assuming *K* ≫ *I*_0_ and hence A ≫ 1. For small time *t*, this approximates the exponential model.

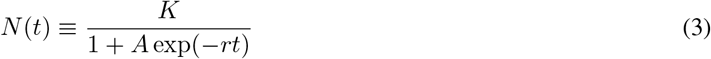

### A.3 SIR Model

Susceptible-Infectious-Recovered (SIR) model is a compartmental model that accounts for the number of susceptibles *S*, the number of infectious *I*, and the number of recovered or deceased (or immune) individuals *R*. Their distributions can be given as following[9] :

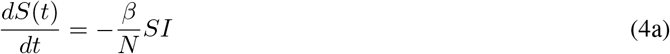

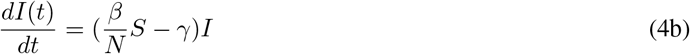

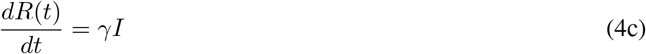

where *β* is the transmission rate, and *γ* is the average recovery rate. Note that here constant *N* is not the population of the region considered but population composed of susceptibles (*S*), infected (*I*) and recovered (*R*).

### A.4 Generalized SEIR (SEIQRDP) Model

The generalized SEIR Model as proposed in Reference[19] includes many more states than SIR model, namely susceptible cases (*S*), insusceptible cases (*P*), exposed cases (*E*), infectious cases (*I*), quarantined cases (*Q*; confirmed and infected), recovered cases (*R*) and deceased cases (*D*). The governing equations for each of them are given in 5. The coefficients *α, β, γ*^∗1^, *δ*^∗1^, *λ*(*t*), *κ*(*t*) represent the protection rate, infection rate, average latent time, average quarantine time, cure rate, and mortality rate, separately. Among these the cure rate *λ*(*t*) = *λ*_0_(1 ∗exp(∗ *λ*_1_*t*)) and mortality rate *κ*(*t*) = *κ*_0_ exp(∗*κ*_1_*t*) are time-dependent, with the former increasing with time while the latter first increases and then rapidly decreases. *λ*_0_, *λ*_1_, *κ*_0_, *κ*_1_ are constant parameters.

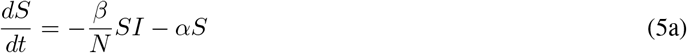

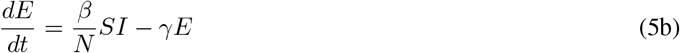

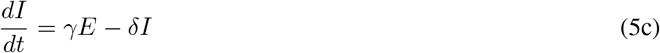

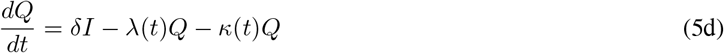

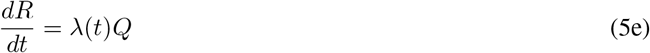

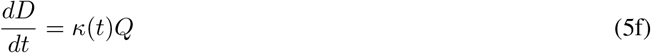

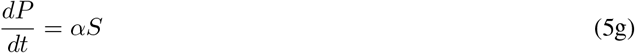

Because of the inclusion of insusceptible cases, here *N* = *S* + *P* + *E* + *I* + *Q* + *R* + *D* represents the total population of a given geographical region unlike the SIR model. The details of this implementation are given in Reference[19].

## Notes

### Competing Interest Statement

The authors have declared no competing interest.

### Funding Statement

No external funding was received.

### Author Declarations

This work does not need any approval of the IRB/oversight body.

